# Novel genetic insights into the roles of amino acids in metabolic dysfunction-associated steatotic liver disease

**DOI:** 10.1101/2024.04.09.24305556

**Authors:** Jun Liu, Yuxuan Chen, Jin Qian, Ri Cui, Ayse Demirkan, Yihu Zheng

**Affiliations:** Nuffield Department of Population Health, University of Oxford, UK; The First School of Medicine, Wenzhou Medical University, China; Department of General Surgery, the First Affiliated Hospital of Wenzhou Medical University, China; School of Pharmaceutical Sciences, Wenzhou Medical University, China; People-centred AI institute & Section of Statistical Multi-Omics, Department of Clinical & Experimental Medicine, School of Biosciences & Medicine, University of Surrey, Guildford, UK

**Author notes:** **Corresponding authors** Jun Liu; Yihu Zheng.

**Keywords:** Metabolic dysfunction-associated steatotic liver disease, amino acids, valine, leucine, alanine, branched-chain amino acids, phenylalanine, Mendelian randomization

## Abstract

**Background:** Previous research has suggested potential links between amino acids and metabolic dysfunction-associated steatotic liver disease (MASLD), but the precise roles of amino acids in MASLD development are not well understood. This study aimed to obtain insights into the relationships between circulating amino acids and MASLD.

**Methods:** Utilizing data from the UK Biobank, we examined the observational associations of ten amino acids with MASLD in a cohort of 72,626 MASLD cases and 128,102 controls. Bi-directional two-sample Mendelian randomization (MR) was conducted using genome-wide association study data to investigate the causal relationships between amino acids and MASLD. Multiple MR methods comprising MR-Egger and MR-PRESSO were applied to assess pleiotropy and heterogeneity, and multivariable MR was conducted to evaluate the impacts of body mass index (BMI) on these associations. Survival analysis assessed the link between baseline amino acid levels and the risk of major outcomes.

**Results:** We identified nine amino acids significantly associated with MASLD in the observational study. The genetic predisposition towards higher leucine (odds ratio (OR) [95% confidence interval (CI)]: 2.1 [1.4, 3.2]), valine (OR [95% CI]: 1.8 [1.3, 2.7]), and alanine (OR [95% CI]: 1.4 [1.1, 1.8]) levels were significantly associated with MASLD. By contrast, the genetic predisposition for increased MASLD risk was significantly associated with phenylalanine (beta = 0.05, *p* = 4.0×10^-4^). Further analysis showed that valine may mediate the association between BMI and MASLD, and may also have an exclusive effect on MASLD in addition to the effect of obesity (beta = 1.3, *p* = 1.9×10^-4^). Elevated phenylalanine levels in MASLD patients were linked with an increased risk of metabolic dysfunction-associated steatohepatitis (MASH), hepatocellular carcinoma, cirrhosis, heart failure, stroke, and mortality.

**Conclusion:** We found genetic associations between circulating branched-chain amino acids, particularly leucine and valine, and MASLD, independent of obesity. Phenylalanine was identified as a potential biomarker for MASLD prognostic complications. These results highlight the importance of amino acid metabolism in MASLD as well as suggest new possibilities for research and therapeutic intervention.

## Introduction

Metabolic dysfunction-associated steatotic liver disease (MASLD), previously known as non-alcoholic fatty liver disease, is a multifactorial condition with a global prevalence of approximately 25% [1]. The progressive form of MASLD, metabolic dysfunction-associated steatohepatitis (MASH), can lead to severe liver complications, and thus MASLD is a leading cause of liver-related mortality. The disease is closely linked to obesity and metabolic syndrome, where it manifests as hepatic insulin resistance and is associated with comorbidities such as type 2 diabetes and cardiovascular diseases [1–2]. Amino acids, particularly branched-chain amino acids (BCAAs), are related to obesity and essential in metabolic regulation. They have been implicated in the pathogenesis of various metabolic disorders. The BCAAs consist of valine, leucine, and isoleucine are essential amino acids obtained from dietary sources. Previous studies have suggested roles for BCAAs in the onset of type 2 diabetes and cardiovascular diseases, and emerging evidence suggests their involvement in MASLD [3–5]. Increased levels of fasting plasma BCAAs are partly mediate the association between MASLD and incident type 2 diabetes [6].

Despite the fact that observational studies have frequently reported higher plasma concentrations of amino acids, particularly BCAAs, in cases of MASLD [7], the nature of their relationship is unclear. Mendelian randomization (MR) uses genetic variations as instrumental variables to investigate potential relationships and their directions, thereby providing an alternative approach that can help reduce the effects of confounding factors and reverse causation. A study based on MR linked MASLD with increased plasma tyrosine levels, suggesting amino acids other than BCAA may also be relevant [8]. Furthermore, MR findings indicate associations between higher levels of alanine and glutamine and the risk of MASLD [9], thereby contributing to our understanding of the metabolic underpinnings of MASLD. However, MR methodology is improved by the day, so as the strength of instrumental variables detected by larger association studies. Additionally, there is a gap in literature as most of the MR research were not supported by individual data or follow-up analysis in addition to the MR results, such as the role of body mass index (BMI) in the association and the effects of biomarkers on severe outcomes related to MASLD. Therefore, further research is needed to clarify these relationships.

In the present study, we aimed to elucidate the genetic underpinnings of the relationships between circulating amino acids and MASLD by employing MR. By utilizing the extensive data in the UK Biobank and advanced MR techniques, we assessed the associations between specific amino acids and the risk of MASLD and major outcomes among patients, as well as the potential mediating role of BMI in these relationships, as BMI is the most important risk factor for MASLD and it is also strongly associated with amino acids. Our findings provide a more definitive understanding of the roles of amino acids in MASLD to potentially inform future therapeutic strategies and contribute to the prevention of this increasingly prevalent disease.

## Methods

### Study population

Our study was conducted based on the UK Biobank, which is a large-scale cohort study that includes over 500,000 participants who were aged between 37 and 73 years during the recruitment period (2006 to 2010). These participants were registered with the UK National Health Service and from 22 assessment centers across England, Wales, and Scotland. Standardized procedures were used for data collection, including a wide range of questionnaires, whole-body magnetic resonance imaging, physical and anthropometric measurements, clinical biomarkers, metabolite measurements, genotype data, and electronic health record linkage. All participants provided electronically signed informed consent, and the study was approved by the North West Multi-centre Research Ethics Committee, Patient Information Advisory Group, and Community Health Index Advisory Group. The current study is part of UK Biobank project 61054. Further details about the UK Biobank are available online.

We excluded participants with non-European ancestry, excessive alcohol consumption (regularly drinking > 60 g/day for males or > 50 g/day for females), patients with other underlying conditions [10], and samples which were not measured by the nuclear magnetic resonance (NMR) spectroscopy metabolic panel at baseline. After applying these exclusion criteria, we retained 200,728 participants for the current study.

### Definitions of MASLD, major outcomes, and covariates

We identified cases of MASLD based on fatty liver index ≥ 60 and presence of at least one of five cardiometabolic criteria reported previously by Rinella ME, et al, at baseline [11]. Other underlying conditions of liver diseases and major outcomes (stroke, heart failure, myocardial infarction, MASH, cirrhosis and hepatocellular carcinoma, and mortality) were defined based on self-reported, hospital inpatient records, or death registration up to November 2022. We considered the earliest documented instance of a condition as the official diagnosis date. Prevalent cases referred to participants who had a recorded diagnosis date on or before their initial assessment, or those who self-reported a condition at that time, and were excluded from the survival analysis. The censor date was set as the earliest date of the first recorded event, the date of death, or the latest recorded date. Alcohol intake was quantified based on a questionnaire about average weekly alcohol consumption. Medication usage was obtained from a touchscreen questionnaire at baseline. The definitions of other covariates, including BMI, blood biochemical biomarkers, fasting time, type 2 diabetes, hypertension, medication information, smoking status, smoking pack-years, education, and physical activity, were presented in a previous study [10].

We utilized the R package *missRanger*, which employs chained random forests, for rapid imputation of missing data necessary to define FLI and MASLD. The imputation process incorporated a range of baseline information, including age, sex, smoking status, pack-years of smoking, frequency of alcohol consumption, physical activity levels as categorized by the International Physical Activity Questionnaire (IPAQ), ethnicity, BMI, education level, blood pressure, and waist-hip ratio. To summarize, the extensive data matrix underwent imputation with up to ten iterations of chained equations and 200 trees, with the process being weighted according to the count of existing non-missing values. During the predictive mean matching phase, three potential non-missing values were chosen as candidates for each missing entry.

### Measurement of amino acids

Baseline amino acids were measured in plasma using the high-throughput ^1^H-NMR metabolomics platform (Nightingale Health, Helsinki, Finland) in a randomly selected subset of 274,124 samples from the UK Biobank [12]. This platform uses a standardized protocol for sample quality control, sample preparation, data storage, and automated spectral analyses, which was previously described in detail [13]. To prepare the data for analysis, we performed natural logarithm transformation of the ten available amino acid variables (i.e., valine, leucine, isoleucine, histidine, glutamine, glycine, tyrosine, alanine, phenylalanine, and total BCAA,) reported by the platform. Metabolite values were rank-based inverse normal transformed before running any analysis.

### Statistical analysis

All analyses were performed using R statistical software (version 4.0.3) and two-tailed tests were conducted.

To investigate the associations between amino acids and MASLD, we performed logistic regression analysis for each separate amino acid. The Cox proportional hazards model was used to assess the associations between baseline amino acids levels and the risk of incident major outcomes during follow-up in MASLD patients. We used three models to adjust for potential confounding factors: model 1 included age, sex, and fasting time; model 2 included the covariates in model 1 as well as common lifestyle factors, including smoking status, number of pack-years of smoking, grams of alcohol consumption per week, education, and physical activities; and model 3 included the covariates in model 2 and BMI. Multiple testing was considered with adjustment of the independent equivalent number of tests by Matrix Spectral Decomposition (MSD) method [14].

For the amino acids that had significant observational associations with MASLD, we further explored their associations by conducting two-sample bi-directional MR using the *TwoSampleMR* R package. The MR analyses were predicated on three assumptions: (1) the genetic variants were associated with exposure; (2) the genetic instruments were not associated with the outcomes through confounding factors; and (3) the genetic instruments did not have direct effects on the outcome, but potentially only through exposure (Figure 1).

**Figure 1.**
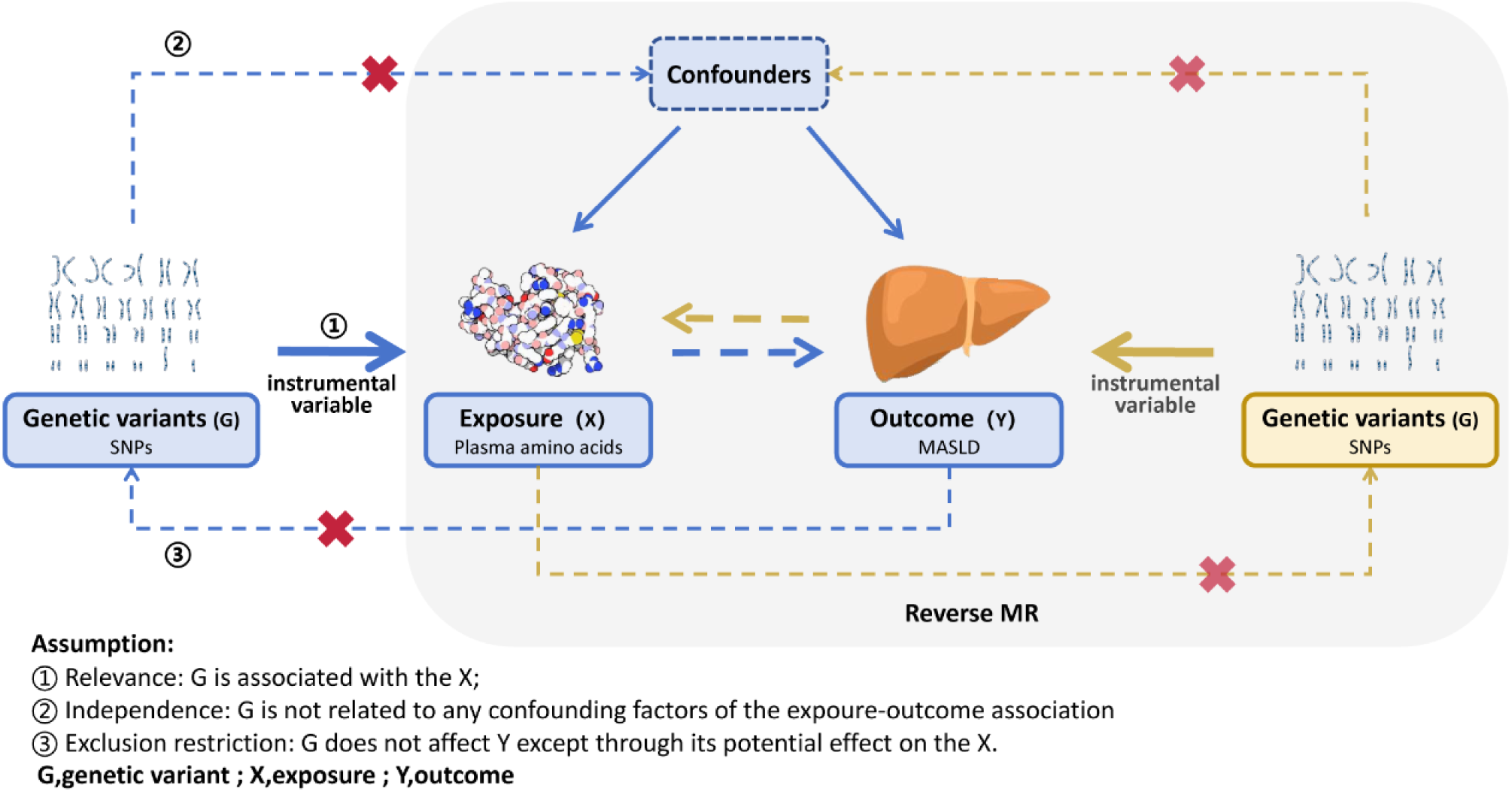
Assumptions of the bi-directional Mendelian randomization study design.

We used data from the largest available genome-wide association study (GWAS) for amino acids from the UK Biobank, which included 115,082 European participants [15]. The GWAS of MASLD was obtained from the study by Ghodsian et al., which included 8,434 MASLD patients and 770,180 controls [16]. Instrumental variables were selected using the independent genetic determinants (*p* < 5×10^-8^) of exposure (either amino acid or MASLD) with a clumping process to exclude single nucleotide polymorphisms (SNPs) in linkage disequilibrium (threshold: R2 > 0.001 and distance < 10,000 kb) [17]. The inverse variance weighted (IVW) method under random effects was used as the primary statisfical method. We also calculated the F statistics for weak instrument bias using the formula: F = [(N – K − 1)/K]*[R^2^/(1 − R^2^)], where N is the sample size, K is the number of IVs, and R^2^ is the proportion of the variability of the exposure explained by IVs.

Sensitivity analysis was performed for the significant MR results with further excluding potentially ambiguous SNPs due to reference strand or palindromic SNPs. Cochran’s Q-test was conducted to assess the heterogeneity across the individual effect estimates derived from each genetic variant. We further tested our hypothesis using other MR methods; weighted median regression [18], MR-Egger regression [19], weighted mode [20], simple mode [20], and MR-Pleiotropy RESidual Sum and Outlier (MR-PRESSO) [21].

For the amino acids potentially on the pathway to MASLD, we further performed multivariable MR to explore the role of BMI in the association of these amino acids and MASLD, using the R package TwoSampleMR with BMI GWAS from Yengo et al., which included 681,275 European participants [22]. The same GWAS sources and pipeline were applied for the MR of BMI and amino acids.

## Results

In the current study, we included 72,626 pure MASLD cases and 128,102 controls. As shown in Table 1, patients with MASLD exhibited increased level metabolic risk factors, including older age, a higher proportion of males, higher BMI, larger waist circumference, higher glucose levels, higher HbA1c levels, higher triglycerides, and higher liver enzymes. Remarkably, these patients had less favorable lifestyles, such as higher rates of smoking and alcohol consumption, lower levels of education, and lower physical activity.

**Table 1.** Characteristics of study population.

| MASLD Variables | Control (n = 128,102) | MASLD (n = 72,676) | p-value |
| --- | --- | --- | --- |
| Age (years), Mean ± SD | 56.21 ± 8.26 | 57.43 ± 7.86 | $< 1 \times 10^{-4}$ |
| Male, n (%) | 41794 (32.63) | 42370 (58.3) | $< 1 \times 10^{-4}$ |
| BMI (kg/m<sup>2</sup>), Mean ± SD | 24.95 ± 2.88 | 31.8 ± 4.58 | $< 1 \times 10^{-4}$ |
| Waist circumference (cm), Mean ± SD | 82.35 ± 8.93 | 102.58 ± 10.19 | $< 1 \times 10^{-4}$ |
| Smoking status, n (%) |  |  |  |
| Never | 81259 (63.43) | 38536 (53.02) | $< 1 \times 10^{-4}$ |
| Previous | 36400 (28.41) | 27040 (37.21) |  |
| Current | 10443 (8.15) | 7100 (9.77) |  |
| Pack years<sup>†</sup>, Median (IQR) | 0.03 (0.03, 0.03) | 0.03 (0.03, 11.5) | $< 1 \times 10^{-4}$ |
| Alcohol grams per week<sup>‡</sup>, Median (IQR) | 44.8 (4, 91.2) | 38.4 (4, 101.6) | 0.029 |
| Education, n (%) |  |  |  |
| College or University degree | 45007 (35.13) | 18946 (26.07) | $< 1 \times 10^{-4}$ |
| A levels AS levels or equivalent | 14645 (11.43) | 7251 (9.98) |  |
| CSEs or equivalent | 6582 (5.14) | 4308 (5.93) |  |
| NVQ or HND or HNC or equivalent | 7173 (5.6) | 6049 (8.32) |  |
| O levels GCSEs or equivalent | 27987 (21.85) | 15359 (21.13) |  |
| Other professional qualifications | 6720 (5.25) | 4151 (5.71) |  |
| None | 19988 (15.6) | 16612 (22.86) |  |
| Physical activity, n (%) |  |  |  |
| High | 55235 (43.12) | 24596 (33.84) | $< 1 \times 10^{-4}$ |
| Low | 20541 (16.03) | 18383 (25.29) |  |
| Moderate | 52326 (40.85) | 29697 (40.86) |  |
| Glucose (mmol/L), Mean ± SD | 4.95 ± 0.79 | 5.32 ± 1.31 | $< 1 \times 10^{-4}$ |
| HbA1c (mmol/mol), Mean ± SD | 35.06 ± 4.67 | 38.24 ± 7.66 | $< 1 \times 10^{-4}$ |
| Anti-diabetes medications, n (%) | 2336 (1.82) | 5898 (8.12) | $< 1 \times 10^{-4}$ |
| T2D, n (%) | 3161 (2.47) | 9014 (12.4) | $< 1 \times 10^{-4}$ |
| Systolic blood pressure (mmHg), Mean ± SD | 134.44 ± 18.67 | 141.49 ± 17.56 | $< 1 \times 10^{-4}$ |
| Diastolic blood pressure (mmHg), Mean ± SD | 79.63 ± 9.74 | 85.01 ± 9.75 | $< 1 \times 10^{-4}$ |
| Anti-hypertensives, n (%) | 20703 (16.16) | 26087 (35.89) | $< 1 \times 10^{-4}$ |
| Hypertension, n (%) | 54640 (44.52) | 49516 (70.33) | $< 1 \times 10^{-4}$ |
| Triglycerides (mmol/L), Median (IQR) | 1.22 (0.92, 1.65) | 2.13 (1.58, 2.89) | $< 1 \times 10^{-4}$ |
| HDL-cholesterol (mmol/L), Mean ± SD | 1.54 ± 0.36 | 1.22 ± 0.28 | $< 1 \times 10^{-4}$ |
| Lipid lowering drugs, n (%) | 20530 (16.03) | 22364 (30.77) | $< 1 \times 10^{-4}$ |
| AST, Median (IQR) | 23.3 (20.2, 27) | 25.9 (22.1, 30.9) | < 1×10<sup>-4</sup> |
| ALT, Median (IQR) | 17.5 (13.97, 22.37) | 25.44 (19.43, 34.26) | < 1×10<sup>-4</sup> |
| GGT, Median (IQR) | 20.5 (15.9, 28) | 36.4 (26.4, 54.4) | < 1×10<sup>-4</sup> |
| CRP, Median (IQR) | 0.98 (0.51, 1.97) | 2.24 (1.19, 4.31) | < 1×10<sup>-4</sup> |

<sup>†</sup>: Never smokers were not considered when calculating pack years. <sup>‡</sup>: Never drinkers were not considered when calculating alcohol grams per week. SD: standard deviation; BMI: body mass index; IQR: interquartile range; AS: Advanced Subsidiary; CES: Certificate of Secondary Education; NVQ: National Vocational Qualification; HND: Higher National Diploma; HNC: Higher National Certificates; GCSE: General Certificate of Secondary Education; HbA1c: Haemoglobin A1c; HDL: high-density lipoprotein; AST: Aspartate aminotransferase; ALT: Alanine aminotransferase; GGT: γ-glutamyl transferase; CRP: c-reactive protein.

### Observational associations between plasma amino acids and MASLD

We compared the levels of ten plasma amino acids between MASLD cases and controls across three models (Table 2). Nine out of the ten amino acids were statistically significantly associated with MASLD across all three models (*p* < 6.25×10^-3^), including six amino acids (valine, leucine, isoleucine, tyrosine, alanine, and phenylalanine) and total BCAA that had positive associations with MASLD, and glutamine and glycine, which had negative associations with MASLD. The effect estimates for these nine amino acids with MASLD were very similar under model 1 and model 2, thereby suggesting that common lifestyle factors had limited effects on their associations. The absolute effect estimates for these nine amino acid variables on MASLD generally decreased after adjustment for BMI on top of model 2 (model 3). The association between histidine and MASLD was significant in three models, but the direction of effect estimates shifted from negative association (beta = –0.34 in model 1 and beta = –0.03 in model2) to positive association (beta = 0.03) after adjusting for BMI, which suggests that their negative associations may be masked by the risk effect of BMI on MASLD.

**Table 2.**
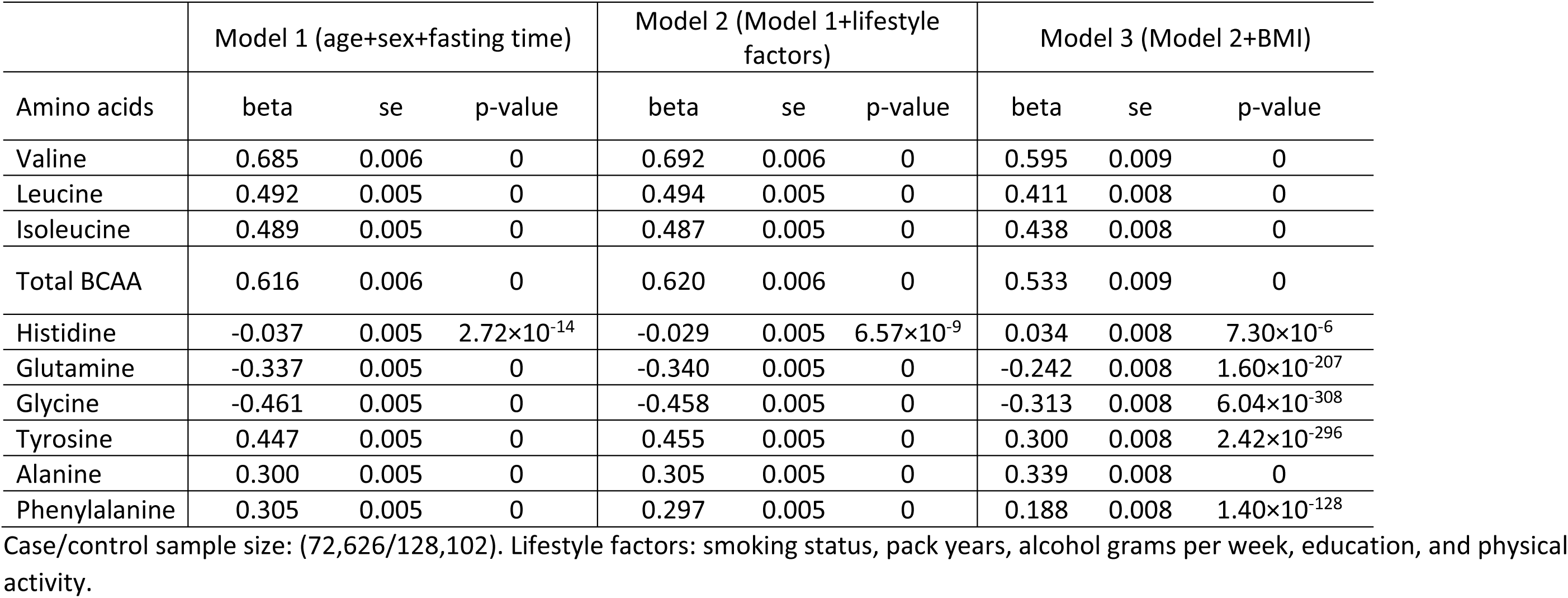
Observational association of plasma amino acids and MASLD.

|  | Model 1 (age+sex+fasting time) |  |  | Model 2 (Model 1+lifestyle factors) |  |  | Model 3 (Model 2+BMI) |  |  |
| --- | --- | --- | --- | --- | --- | --- | --- | --- | --- |
| Amino acids | beta | se | p-value | beta | se | p-value | beta | se | p-value |
| Valine | 0.685 | 0.006 | 0 | 0.692 | 0.006 | 0 | 0.595 | 0.009 | 0 |
| Leucine | 0.492 | 0.005 | 0 | 0.494 | 0.005 | 0 | 0.411 | 0.008 | 0 |
| Isoleucine | 0.489 | 0.005 | 0 | 0.487 | 0.005 | 0 | 0.438 | 0.008 | 0 |
| Total BCAA | 0.616 | 0.006 | 0 | 0.620 | 0.006 | 0 | 0.533 | 0.009 | 0 |
| Histidine | -0.037 | 0.005 | $2.72 \times 10^{-14}$ | -0.029 | 0.005 | $6.57 \times 10^{-9}$ | 0.034 | 0.008 | $7.30 \times 10^{-6}$ |
| Glutamine | -0.337 | 0.005 | 0 | -0.340 | 0.005 | 0 | -0.242 | 0.008 | $1.60 \times 10^{-207}$ |
| Glycine | -0.461 | 0.005 | 0 | -0.458 | 0.005 | 0 | -0.313 | 0.008 | $6.04 \times 10^{-308}$ |
| Tyrosine | 0.447 | 0.005 | 0 | 0.455 | 0.005 | 0 | 0.300 | 0.008 | $2.42 \times 10^{-296}$ |
| Alanine | 0.300 | 0.005 | 0 | 0.305 | 0.005 | 0 | 0.339 | 0.008 | 0 |
| Phenylalanine | 0.305 | 0.005 | 0 | 0.297 | 0.005 | 0 | 0.188 | 0.008 | $1.40 \times 10^{-128}$ |

Case/control sample size: (72,626/128,102). Lifestyle factors: smoking status, pack years, alcohol grams per week, education, and physical activity.

### Bi-directional MR of amino acids and MASLD

For the nine amino acids that had robust significant associations with MASLD, we performed two-sample bi-directional MR based on the previously published GWAS of MASLD and the GWAS of the UK Biobank [15–16] (Figure 2). The number of genetic loci selected in the instrumental variables for amino acids ranged from seven for isoleucine and phenylalanine to as many as 35 for glycine, with F statistics ranging from 62.2 for leucine to 603.9 for glycine. We found that the genetic predisposition towards increased levels of leucine (odds ratio (OR) [95% confidence interval (CI)]: 2.1 [1.4, 3.2]), valine (OR [95% CI]: 1.8 [1.3, 2.7]), and alanine (OR [95% CI]: 1.4 [1.1, 1.8]) were statisfically significantly associated with increased risk of MASLD (*p* < 3.6×10^-3^).

**Figure 2.**
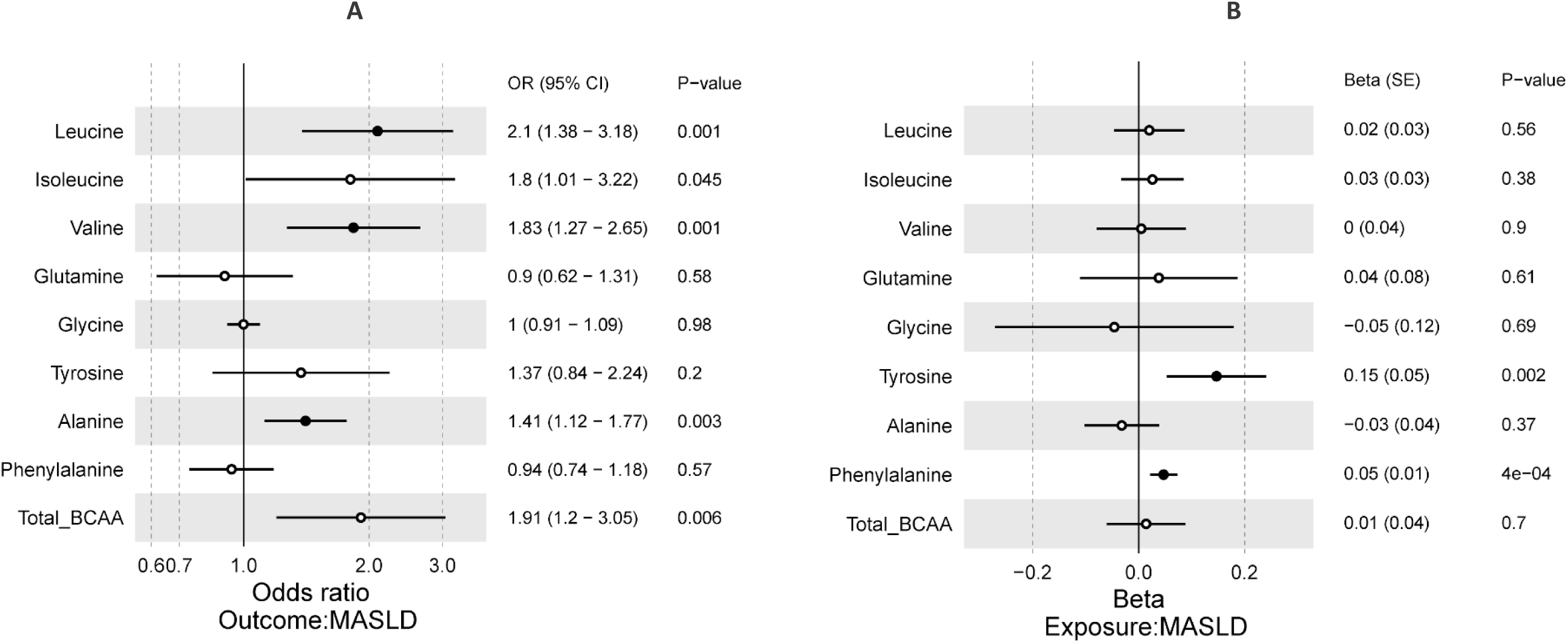
Forest p lots showing the effects of amino acids on MASLD (A) and MASLD on amino acids (B) using the inverse variance-weighted method. CI: Confidence interval. SE: Standard error. Solid dots indicate significance with *p* < 6.25×10^-3^. Hollow circles indicate non-significance with *p* ≥6.25×10^-3^.

Four genetic loci were selected as the instrumental variables for MASLD, i.e., rs28601761, rs3747207, rs429358, and rs73001065, which had F statistics of 13.4. The F statistics for MASLD were not as strong as those for amino acids, but we still detected statistically significant differences between the genetic predisposition toward MASLD and increased levels of tyrosine (beta = 0.15, *p* = 2.0×10^-3^) as well as phenylalanine (beta = 0.047, *p* = 4.0×10^-4^) (*p* < 3.6×10^-3^).

### Sensitivity analysis based on significant MR findings

For the significant MR analysis results, including three amino acids (valine, leucine, and alanine) relative to MASLD and MASLD relative to two amino acids (phenylalanine and tyrosine), we explored the potential heterogeneity and pleiotropic effect in MR (Figure 3) using additional MR methods. Significant association of between genetic predisposition to elevated valine, leucine, and alanine were confirmed by the weighted mean method, while for the other methods significance varied with different assumptions while directions of the effect estimates were stable (Figure 3A). On the other hand, the effect of MASLD on amino acids phenylalanine and tyrosine was significant for all methods tested, except MR-Egger (Figure 3B). Sensitivity analysis excluding potentially ambiguous and/or palindromic SNPs in the instrumental variables did not affect the significance of the MR analysis results, except the association between MASLD and tyrosine shifted to nominal significance (*p* = 0.022) when we excluded rs28601761 (Figure 3C-D).

**Figure 3.**
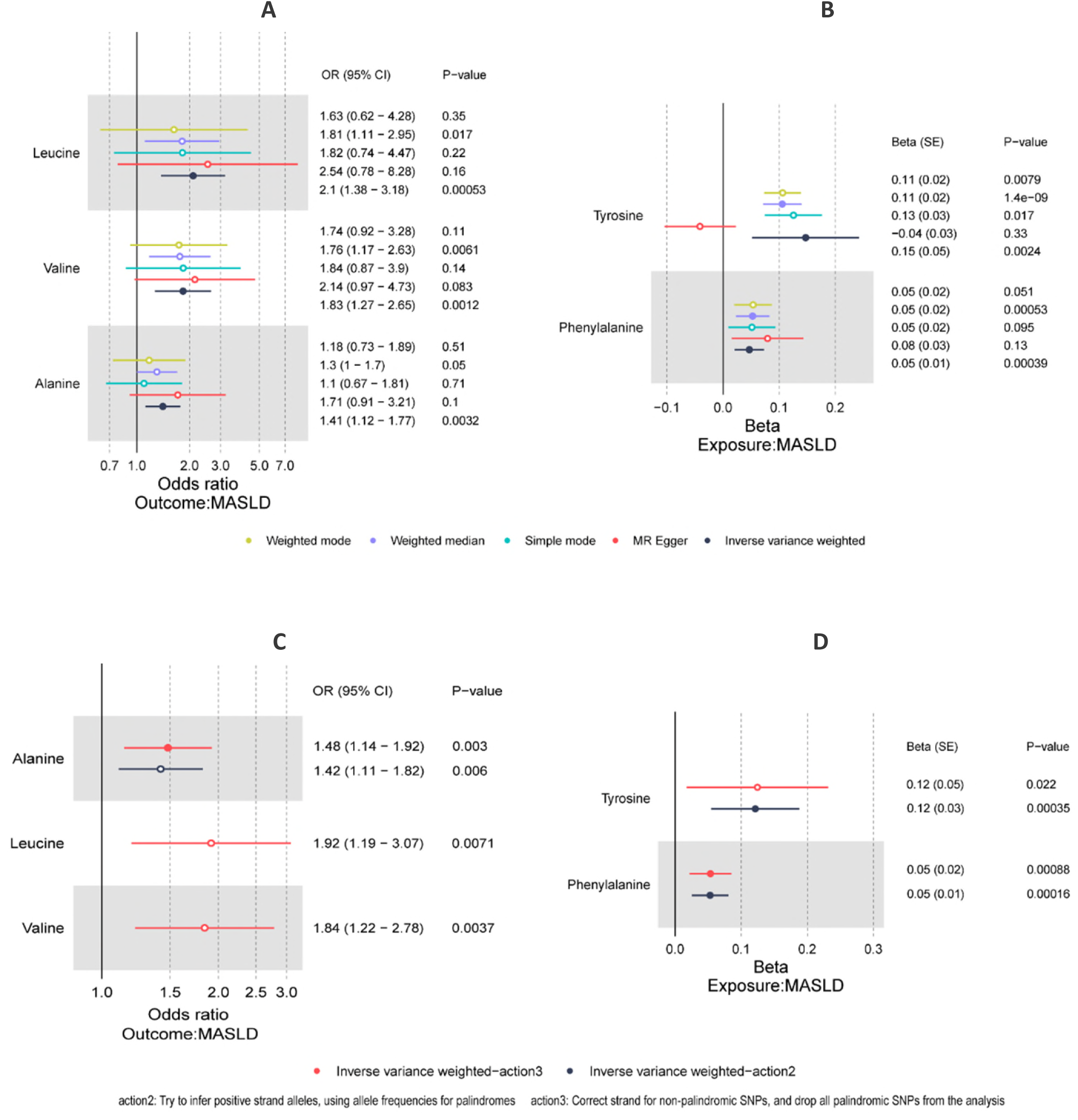
Forest plot showing the effects of leucine, valine, alanine on MASLD and MASLD on tyrosine and phenylalanine. (A-B) Using different Mendelian randomization analyses; (C-D) Using the inverse variance-weighted method with stricter conditions. CI: Confidence interval. SE: Standard error. Solid dots indicate significance with *p* < 6.25×10^-3^. Hollow circles indicate non-significance with *p* ≥ 6.25×10^-3^.

Furthermore, Cochran’s Q-test demonstrated significant heterogeneity among the SNPs included in the instrumental variables for alanine with MASLD (*p* = 5.0×10^-3^) and in the instrumental variables for MASLD with tyrosine (*p* = 8.36×10^-9^; *p* < 0.01). MR-PRESSO was also applied as another method to detect heterogeneity among the SNPs used in the instrumental variables and it consistently detected significant heterogeneity through the global test for alanine with MASLD (*p* = 4.0×10^-3^, *p* < 0.01). The distortion test detected no difference in the MR results before and after excluding outliers for the link between alanine and MASLD (*p* > 0.01). Moreover, no significant pleiotropic effect was detected by MR-Egger regression (*p* > 0.01). The MR results for leucine, valine, and phenylalanine were robust according to heterogeneity and pleiotropy tests.

### Role of BMI in the associations between amino acids and MASLD

BMI is the most important risk factor for MASLD and it is also strongly associated with amino acids, so it was essential to explore its role in the associations between amino acids and MASLD. First, we investigated the potential role of BMI in the associations of the three amino acids (leucine, valine, and alanine) and MASLD by MR (Table 3). We found that the genetic predisposition towards higher BMI was strongly associated with both BCAAs, i.e., leucine and valine (beta = 0.19, *p* = 1.84×10^-27^ for leucine; beta = 0.23, *p* = 1.36×10^-37^ for valine), through IVW. MR-PRESSO showed that the heterogeneity among the instrumental variables did not affect their significance (*p* > 0.05) and no pleiotropic effect was detected. The association of genetic predisposition towards higher BMI and alanine was borderline significant (*p* = 0.02, *p* < 0.017).

**Table 3.** IVW-MR results and sensitivity analysis of the association between BMI (exposure) and plasma amino acids associated to MASLD (outcome)

| Outcome | beta | se | p-value | Pleiotropy<br>(p-value) | MR-PRESSO<br>(p-value) |
| --- | --- | --- | --- | --- | --- |
| Leucine | 0.189 | 0.017 | $1.84 \times 10^{-27}$ | 0.446 | 0.63 |
| Valine | 0.232 | 0.018 | $1.36 \times 10^{-37}$ | 0.725 | 0.491 |
| Alanine | 0.041 | 0.018 | 0.02 | 0.318 | 0.768 |

After considering the effect of BMI on the associations of leucine, valine, and alanine with MASLD by multivariable MR (Table 4), a significant association remained for valine (beta = 1.28, *p* = 1.9×10^-4^), which suggested potential pathways from valine to MASLD independent of the obesity effect. A high beta value was also obtained for leucine (beta = 2.9). No significance was detected for alanine and MASLD after adjusting for the effect of BMI (*p* = 0.31), thereby suggesting that the association between alanine and MASLD was likely to be fully mediated by the effect of obesity.

**Table. 4.** Multivariable MR results of the association between plasma amino acids (exposure) and MASLD (outcome), mediating by BMI effect.

|  | BMI |  |  |  | Amino acids |  |  |  |
| --- | --- | --- | --- | --- | --- | --- | --- | --- |
| exposure | SNP number | beta | se | p-value | SNP number | beta | se | p-value |
| Leucine | 479 | 0.340 | 0.061 | $2.35 \times 10^{-8}$ | 2 | 2.904 | - | - |
| Alanine | 469 | 0.442 | 0.056 | $5.33 \times 10^{-15}$ | 4 | 0.581 | 0.576 | 0.31 |
| Valine | 475 | 0.295 | 0.062 | $1.78 \times 10^{-6}$ | 3 | 1.278 | 0.342 | $1.9 \times 10^{-4}$ |

### Roles of phenylalanine and tyrosine in major outcomes of MASLD patients

Phenylalanine and tyrosine were found to be associated with the genetic predisposition toward MASLD, and thus the alterations in them may result to the progression of MASLD. Therefore, we further explored their associations with the incidence of major outcomes in MASLD patients (Figure 4). The results showed that baseline phenylalanine was significantly associated with increased risk of MASH (hazard ratio (HR) [95% CI]: 1.3 [1.16–1.45]), hepatocellular carcinoma (HR [95% CI]: 1.56 [1.32–1.85]), cirrhosis (HR [95% CI]: 1.14 [1.08–1.2]), heart failure (HR [95 %CI]: 1.13 [1.09–1.14]), stroke (HR [95% CI]: 1.09 [1.04–1.14]), and mortality (HR [95% CI]: 1.06 [1.04–1.09]), and not affected by lifestyle factors or BMI (*p* < 7.1×10^-3^). The associations of tyrosine and increased risk of liver related outcomes with MASH (HR [95% CI]: 1.71 [1.52–1.92]), hepatocellular carcinoma (HR [95% CI]: 2.04 [1.73–2.42]), and cirrhosis (HR [95% CI]: 1.2 [1.14–1.27]) were consistent across the adjusted models, but not associated with any cardiovascular events or mortality after accounting for BMI.

**Figure 4.**
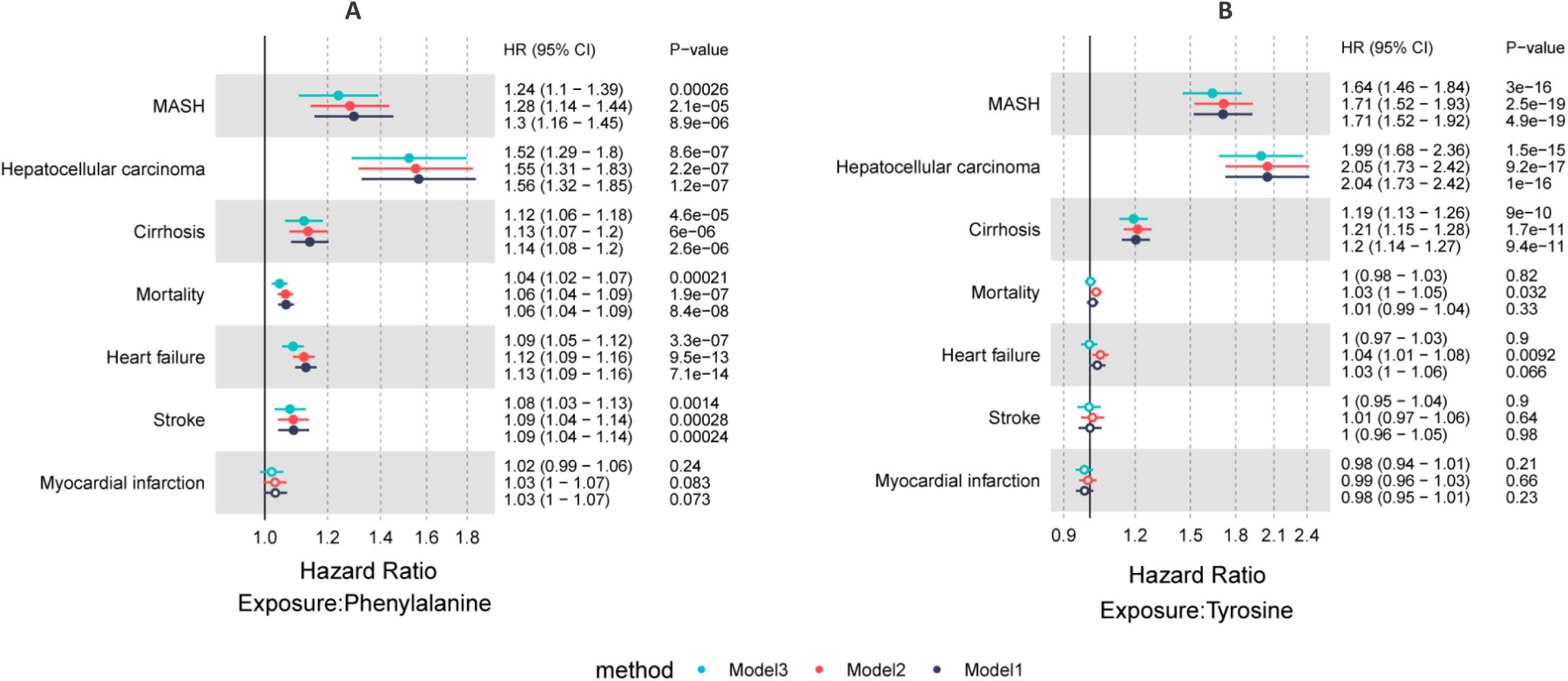
Forest plot showing the associations of phenylalanine (A) and tyrosine (B) with incidence of major outcomes in MASLD patients. CI: Confidence interval. Solid dots indicate significance with *p* < 7.1×10^-3^. Hollow circles indicate non-significance with *p* ≥ 7.1×10^-3^.

## Discussion

In the present study, for the first time, we established a robust genetic link between valine, leucine, and phenylalanine with MASLD using a MR approach. We found that valine and leucine may mediate the relationship between BMI and MASLD but it could also be involved in unique pathways leading to MASLD in addition to the effect of obesity, thereby suggesting that BCAAs could potentially serve as targets for MASLD prevention or treatment. In addition, we identified phenylalanine as a potential novel outcome of MASLD, which could be developed as an early detection biomarker for severe liver and cardiovascular outcomes in MASLD patients.

Following our observational findings which indicated a strong correlation between various amino acids and MASLD, we provided genetic evidence of associations between genetic predisposition of leucine, valine, and alanine with MASLD, and between genetic predisposition of MASLD with tyrosine and phenylalanine. Our results validated previous findings by Zhao et al. [9], who reported that higher plasma alanine levels alter a higher risk of MASLD by MR approach. In addition to that, we obtained further evidence that their association is likely to be mediated by obesity as adjustment for BMI by multivariable MR resulted in a non-significant association (*p* = 0.31). Moreover, our demonstration of an association between MASLD with tyrosine is consistent with previous findings by Gobeil et al. [8] who used different data sources for the MR analysis. Moreover, our results also provide extra information about the effect of tyrosine on the risk of incident liver outcomes (MASH, hepatocellular carcinoma, and cirrhosis) in MASLD patients, indicating that as a result of MASLD development, alterations in tyrosine may play an important role in the liver complications of MASLD. Further MR analyses of tyrosine and liver outcomes in MASLD patients, which are not yet available, could provide more evidence for the observational prospective cohort.

Notably, the associations between leucine and valine with MASLD and the association of MASLD with phenylalanine obtained by the MR approach were determined for the first time with robust results through various sensitivity analyses. Moreover, we also detected a complex interaction between BMI, valine, and MASLD. BMI led to the incidence of MASLD with a significant mediating effect from valine and leucine, and valine also had an exclusive effect on MASLD other than through obesity. These findings suggest that BCAAs, especially valine, could have important roles in the further development of prevention and treatment strategies for MASLD. Currently, the mechanism that allows plasma BCAAs to cause MASLD is not well understood. However, it has been shown that overconsumption of valine triggers MASLD in laying hens by stimulating fatty acid synthesis in the liver [23]. Experimental studies have shown that elevated BCAAs can damage the oxidation of carbon substrates through the tricarboxylic acid cycle (TCA cycle) to result in mitochondrial dysfunction and participation in MASLD [24]. In addition, leucine promotes MASLD by modulating AMP-activated protein kinase through myostatin to lead to hepatocyte triglyceride accumulation [25]. Moreover, aminoacylation, as a novel amino acid modification, allows BCAAs to exert their regulatory roles in cellular functions through the effect of lysine aminoacylation on specific substrate proteins catalyzed by aminoacyl-tRNA synthetase [26]. In addition, many factors can affect the concentrations of plasma BCAAs, including exogenous BCAA uptake, adipose tissue, and skeletal muscle, which may be a part of personalized therapeutic routes for MASLD management involving BCAAs.

In addition to the important roles of BCAAs in the pathway to MASLD, it should be noted that we identified phenylalanine as a new biomarker for MASLD. Previously, the correlation between non-obese individuals with MASLD and phenylalanine levels was discovered using metabolomics approaches [27]. However, no clinical data with a large sample size are available to support this correlation and causal inferences were not found by MR analysis. In the present study, for the first time, we found that alterations in blood phenylalanine levels were associated with the genetic predisposition toward MASLD. Moreover, we provided evidence based on prospective cohort data that the baseline phenylalanine level in MASLD patients may be an early biomarker for liver and cardiovascular major outcomes and mortality. Our results are consistent with those obtained in previous clinical studies. The plasma phenylalanine increased according to the severity of the disease from steatosis to non-alcoholic steatohepatitis compared with normal control [28–29]. Moreover, in a prospective cohort study, the plasma phenylalanine concentration correlated with hepatocellular carcinoma occurrence in liver cirrhotic patients after following for the next 3 years [30].

The present study also had some limitations. First, the GWAS summary statistics for MASLD and amino acids involved overlapping participants, but the overlap for MASLD cases was only 19.7% (1,644 in 8,434), which was unlikely to have affected our major findings [31]. Second, in case of SNP missingness, we did not opt for replacing them with proxy SNPs for the instrumental variables of exposures, which may have reduced the explained variance for the instrumental variables on exposure. However, this only increased the likelihood of false-negative findings and would not have affected our significant results. Third, we only included ten amino acids in the study. However, individual data were available for these amino acids to support their observational associations with MASLD and MASLD complications. Finally, more tissue-based studies are required to understand the mechanisms that underlie the relationships between plasma BCAAs and MASLD.

In conclusion, we established a link between circulating BCAAs and MASLD using a MR approach, thereby suggesting that BCAAs could potentially serve as targets for the prevention or treatment of MASLD. We also highlighted the complex interactions between BMI, BCAAs, and MASLD, and the important roles of BCAAs in the development of prevention and treatment strategies for MASLD. Furthermore, we identified alteration in phenylalanine as a novel potential outcome of MASLD, which could be developed as a biomarker for the early detection of MASLD and predicting the incidence of major outcomes in MASLD patients. Our study significantly contributes to understanding the interplay between amino acids and MASLD to provide a foundation for future research and the potential development of therapeutic strategies. Further tissue-based studies are required to elucidate the mechanisms that underlie their relationships.

## Author contributions

J.L. and Y.Z. conceived and designed the current study. J.L., Y.C., and J.Q. performed analyses. J.L, Y.C., and Y.Z. prepared the manuscript. R.C, A.D. and Y.Z. reviewed and edited the manuscript. All authors read and approved the manuscript.

## Data availability

UK Biobank data are publicly available to bona tide researchers upon application at http://www.ukbiobank.ac.uk/using-the-resource/. Publicly available summary statistics are obtained from https://gwas.mrcieu.ac.uk/ andhttps://www.ebi.ac.uk/gwas/. Other sources of data or web sources were clarified in the methods.

## Acknowledgement

This research was conducted using data from UK Biobank, a major biomedical database (https://www.ukbiobank.ac.uk/) via application no. 61054. We thank the participants, contributors, clinicians, and researchers for making data available for this study. J.L. is supported by a Novo Nordisk Postdoctoral Fellowship Programme run in partnership with the University of Oxford. Y. Z is supported by Discipline Cluster of Oncology, Wenzhou Medical University, China (No.z2-2023024).

